# Device-measured movement behaviours and cancer incidence in a population sample of UK adults: Dual 24-hour analyses of postural and intensity compositions

**DOI:** 10.1101/2025.10.02.25337148

**Authors:** John J Mitchell, Raaj Biswas, Nicholas Koemel, Matthew Ahmadi, Joanna M Blodgett, Karen Canfell, I-Min Lee, Christine Friedenreich, Armando Teixeira-Pinto, Peter A. Cistulli, Dorothea Dumuid, Anthony D. Okely, Abigail Fisher, Mark Hamer, Emmanuel Stamatakis

## Abstract

**IMPORTANCE:** Insufficient physical activity (PA) and excessive sedentary behaviour is associated with several cancers. Personalised approaches to increasing healthy movement behaviours and reducing unhealthy behaviours may be more effective than a one-size-fits-all approach. Exploring doses, posture allocation and intensities across the 24-hour spectrum of behaviours are important to develop broader guidelines.

**OBJECTIVE:** Using a novel dual-compositional approach, we assessed how time reallocation between 24-hour daily movement (postures and intensity bands) and sleep are associated with PA-related cancer incidence (a composite of 13 sites linked with physical inactivity).

**DESIGN, SETTING, AND PARTICIPANTS:** Prospective analysis of adults from the UK Biobank accelerometry subsample, followed-up by health linkage.

**EXPOSURES:** Participant’s daily movement was classified into two 24-hour compositions based on intensity and posture. Composition 1 (posture-focused): sleep duration, sedentary behaviour (SB), standing, moving at any intensity. Composition 2 (intensity-focused): sleep duration, sedentary time (ST), light PA (LPA), moderate PA (MPA), and vigorous PA (VPA). Secondary analysis combined VPA and MPA as moderate-to-vigorous PA (MVPA).

**MAIN OUTCOMES AND MEASURES:** PA-related cancer diagnoses were captured from health linkage. Cox-proportional hazards models were adjusted for age, sex, education, smoking, alcohol, diet, parental cancer history, cardiovascular disease and medication use.

**RESULTS:** Analyses included 59,218 (55% female) participants (mean [SD] age: 61.7 [7.8] years (y)), with a median follow-up of 8.0y [IQR: 7.4-8.5y; 464,640 person years] with 2,385 (4%) incident cancer events. Even among the average, active participant, replacing any behaviour with moving was associated with lower cancer risk, e.g. theoretically replacing 15 min of sleep or SB with 15 min of moving was associated with hazard ratios (HR) of 0.98 (95% Confidence Interval (95%CI): 0.97-0.99) and 0.98 (95%CI: 0.97-0.99), respectively. Similar risk reduction required 30 min of additional standing in place of sleep or ST. Regarding intensity, replacing even 3 min of any other behaviour with equivalent VPA was associated with lower risk (e.g. LPA HR: 0.96 (95%CI: 0.93-0.99); MPA HR: 0.96 (95%CI: 0.93-0.99). Greater MVPA too, in place of any behaviour, was robustly associated with lower risk.

**CONCLUSIONS AND RELEVANCE:** Moving at any intensity, in place of other postures, was associated with reduced risk of incident cancer. VPA, or MVPA in place of other movement intensities, were most strongly associated with lower cancer risk.

**Key points:** *Question:* How may reallocations of time between movement behaviours (postures and intensities) change cancer risk?

*Findings:* Among 59,218 middle-aged UK Biobank participants, 2,385 individuals developed cancer at any of 13 sites linked to physical activity (PA) over 8 years. Even among the average, active participant, replacing 15 minutes of standing, sedentary behaviour (SB) or sleep with moving, or 90+ minutes SB or sleep with standing, was associated with small (2-3%) cancer risk reduction, while even 3 minutes vigorous PA in place of light PA, sedentary time or sleep was associated with 1-7% lower risk.

*Meaning:* Small changes to daily movement (posture/intensity) may reduce cancer risk.

## INTRODUCTION

Cancer is a leading cause of mortality globally. In 2022, there were an estimated 20 million new cancer diagnoses and 9.7 million deaths attributed to cancer^1^. The global incidence of cancer is estimated to reach 34 million new cases in 2070^2^. Recent consensus statements on cancer prevention^4–9^ have focussed on lifestyle behaviours, finding physical inactivity^10,11^ among the most modifiable of risk factors and a target for intervention. Regular engagement in leisure-time physical activity (LTPA) has been previously associated with an 8-25% reduction in cancer risk across a wide range of cancer sites^3, 4^. Nevertheless, physical activity (PA) is only one facet of the 24-hour (h) daily movement spectrum, alongside sleep and sedentary behaviour (SB)^5^, and also encompasses a spectrum of postures and movement intensities. Given the finite nature of time use, it is crucial for public health guidance to understand which behaviours should be a focus of behaviour change interventions for cancer risk reduction. Studies have also reported an adverse relationship between SB, and cancer, independent of PA^6–8^, while the literature focusing on sleep is largely inconclusive^9^. Despite these independent lines of evidence, the field has increasingly acknowledged the interplay between all daily movement behaviours, including SB and sleep for health and longevity^10–12^.

The World Health Organization (WHO) recommends 150-300 minutes of moderate-intensity PA, or 75 minutes of vigorous-intensity PA, weekly^13, 14^. There is however a growing uptake of guidelines for the 24-h movement behaviour (PA, SB and sleep) continuum as this approach may provide a range of options for modifying behaviour to achieve health benefit, instead of a siloed or one-size-fits-all approach^11, 15^. The 24-h movement continuum may be characterised according to many dimensions, including both posture and movement intensity, which can be captured through wearable devices^16, 17^. Advances in wearable algorithms now permits more nuanced exploration of movement behaviours (e.g. posture, intensity, type, duration, etc.) beyond those captured by self-reported questionnaires and can help address calls for evidence on the role of different movement dimensions, including postural allocation for cancer risk^18–20^.When combined with compositional data analysis, which can model all movement in the 24-h day simultaneously, it is possible to establish the relative importance of each behaviour in the context of all others, and to estimate the health effects of changes to daily behaviours by, for example, replacing light or moderate intensity activities with more vigorous activities, or replacing sedentary time with standing or moving^21^.

Despite their methodologic strengths, prior studies that have used a compositional approach typically examine only one dimension of movement behaviour (typically intensity), resulting in a narrow range of possible recommendations^10, 22^. To study two distinct, but clinically-meaningful dimensions of movement behaviour (posture and intensity), and preserve the high-resolution made possible by highly-specific algorithms, it is necessary to extend this to a dual-approach; separately modelling two independent compositions of movement behaviour, sedentary time and sleep in relation to cancer risk.

Using a comprehensive dual 24-h compositional approach and the largest wearables-based data resource to date, this study aimed to quantify the theoretical effects of reallocating time between movement behaviours comprising postures (SB, standing, moving) and intensities (sedentary time (ST), and light, moderate and vigorous intensity PA), and sleep duration on cancer risk.

## METHODS

### Study sample

Participants were drawn from the UK Biobank Study, a prospective cohort study involving 502,629 individuals within the UK, aged 40-69 years (y) at the time of enrolment in 2006 to 2010. Participants provided written informed consent before undertaking a range of physical examinations by trained practitioners and completing detailed health and lifestyle questionnaires.

### Measurement of posture, physical activity, and sleep

The UK Biobank (UKBB) wrist accelerometry sub-study^23^, which received ethical approved from the UK National Research Ethics Service (No. 11/NW/0382), recruited 103,684 individuals from June 2013 to 2015. Sample derivation is displayed in Figure S1. Participants from the UKBB accelerometry sub-study wore an Axivity AX3 accelerometer device (Axivity, Newcastle, UK) on their dominant wrist for 7 days^23^. Devices were initialised to collect movement data with a sampling frequency of 100Hz and a dynamic range between ±8g. A valid day was defined as a minimum of 16 hours of wear time^24^. Participants were included if they had a minimum of three days, including at least one weekend day. Monitors were calibrated^25^, corrected for orientation^26^, and non-wear detected using standard procedures in UKBB^24, 27^. The sleep duration algorithm relies on the absence of change in device tilt angle and was developed and validated in 3,752 British participants with sleep diary data and 28 patients with polysomnography data^28^ (further details are provided in supplementary text S1). Two further activity and posture classifiers, validated in laboratory and free-living conditions^29, 30^ enabled all waking time to be characterised into compositions of intensity, and postures. The details of intensity and posture schemes are outlined below and in supplementary text S1.

To facilitate a comprehensive examination of the theoretical replacement effects of movement behaviour sitting and sleep on cancer risk, we utilised a novel dual-composition approach^31^. This approach enabled investigation of the importance of these related but clinically-distinct dimensions of posture and intensity. The two compositions were defined as follows:

### Composition 1: Posture (4-parts)

The activity classification scheme uses features derived from the raw acceleration signals to classify waking-periods as sitting, standing (both still and with subtle, utilitarian movement), or walking/running in 60-second (s) windows^29, 30^, using a Random-forest classifier with a Hidden Markov Model^32^. Under free-living conditions, the classifier achieved balanced accuracy (average of sensitivity and specificity) of 88% for SB and 80% for standing^30^. Composition 1 comprised the following four 24-h cycle behaviours: sleeping, SB, standing, and moving. Further details of the algorithms are presented in supplementary text S1.

### Composition 2: Intensity (5-parts)

Physical activity intensity was classified with a previously adopted two-level Random Forest algorithm^16, 24^. The first level classifies activity type (moving activities, standing utilitarian movements (semi-stationary) and SB, in contiguous 10s bins. The second level further categorizes activities into intensity bands. Moving activities are then grouped by their acceleration into light PA, moderate PA, and vigorous PA. The intensity composition therefore encompassed five behaviours: sleep, SB, light PA (LPA), moderate PA (MPA) and vigorous PA (VPA) in minutes (min). Further details are presented in supplementary text S1. A secondary intensity composition involved collapsing MPA and VPA to MVPA, producing a 4-part composition.

For both compositions 1 and 2 described above, the average daily time in each component across the week was normalised to sum to 24 hours^33^ (actual averaged wear-time across valid days was 23h 26m). Finally, to handle left-censored data (meaningful movement at intensities below the minimum detection threshold of the device), a log-ratio expectation maximisation algorithm was utilised to multiplicatively replace rounded zeroes^34, 35^.

### Physical activity-related Cancer Incidence

We used a single composite outcome comprising incidence of cancer at any one of 13-sites shown to be associated with leisure-time PA^3, 36^. This PA-related cancer composite variable encompassed bladder, colon, endometrial, gastric cardia, head and neck, kidney, liver, breast, lung, rectal cancer, myeloma, oesophageal adenocarcinoma and myeloid leukaemia. Further details are provided in table S1. Incidence and date of diagnosis was derived via linkage with the National Health Service (NHS) Digital records for participants based in England and Wales, while the NHS Central Register was used to identify cancer events in participants in Scotland. Inpatient hospitalisation data were provided by either the Hospital Episode Statistics for England, the Patient Episode Database for Wales, or the Scottish Morbidity Record for Scotland. Cancer data linkage was obtained through national cancer registries. For England and Wales, cancer diagnosis data were followed-up through to 31 December 2020 and 31 December 2016 respectively^37^. For Scotland, cancer diagnosis data were followed-up through 30 November 2021 and provided by the National Records of Scotland. Participants with any history of cancer (n=7633) were excluded from the sample.

### Covariates

Covariates were selected *a priori* based on previous literature examining the association of device-measured PA and cancer incidence^3, 36 36^. Covariates included age (in years), sex at birth (male vs female), ethnicity (white vs any other ethnic background) and maximal level of education (College/University (attained at >18y; A/AS level (attained at 18y); O levels (attained at 16y); NVQ/HND/HNC (typically attained at >16y); CSE (attained at 16y); other). Health factors included prevalence of CVD, captured by self-report or hospital admission (yes/no), medication use (taking antihypertensives, anti-hypercholesterolaemia or insulin) and family history of cancer (yes/no). Lifestyle factors included smoking status (never, past, current), diet quality (number of daily fruit/vegetable servings) alcohol consumption history (never, ex-drinker, current drinker within guidelines (<15 weekly units), current drinker above UK guideline levels (≥15 weekly units)). Further derivation of each covariate is provided in table S1. Adiposity measures were not included in the adjusted models due to their hypothesised mediating role between 24-hr movement behaviour and cancer risk which would lead to model overadjustment^19^.

### Analyses

Primary analyses involved an established isometric-log ratio (ILR)-coordinate compositional approach^21, 33, 38, 39^, Cox proportional hazards models. This statistical method examines the association of each behaviour, relative to all others, with the outcome. Further details on the compositional modelling procedure are provided in supplementary text S2. Fine-Gray sub-distribution method was used to account for the competing risk of mortality from non-PA-related cancer deaths during the follow-up window. The proportional hazards assumption was examined by inspecting Schoenfeld residuals for each fitted model, and no violations were noted. We conducted two sets of analyses, the first using the 4-part composition defined by posture; the second using the 5-part composition defined by intensity. Each component was examined relative to all others in the composition, in separate models until all possible combinations had been studied. For clinical relevance, secondary analysis repeated the primary models with the four-part composition involving MVPA in place of MPA and VPA, and also using a VPA:MPA ratio-variable to better understand the relative importance of the VPA and MPA within daily MVPA.

Minute-by-minute isotemporal substitution then assessed the difference in absolute risk from theoretically reallocating time from one behaviour (those significantly associated with cancer in the fully-adjusted models), to another, whilst holding all other behaviours fixed at the sample average. To provide insights relevant to translation and interventions^36, 40^, estimated change in risk was reported for reallocation increments of 3,5,10,15,30,45,60 and 90m. The chosen increments were selected to be informative for the full range of behaviours, while ensuring modelling within the limits of the data (i.e. 3 min exchange from VPA may be extrapolative given the low sample average VPA).

### Sensitivity analyses

Any individuals with cancer events occurring in the first and second year of follow-up, were sequentially removed, to assess the possibility of reverse causation (i.e. existing disease driving low PA, high SB, etc)^36^. Thirdly, E-values were calculated to quantify the minimum strength of association which an unmeasured confounder would require with both the exposure and outcome to nullify any association^41^. Models were re-fitted with cancer at all sites due to limited consensus surrounding PA-Cancer mechanisms. Finally, we compared demographics of the analytical sample to the excluded sample to assess possible selection bias.

## RESULTS

### Participant demographics

A total of 59,218 participants (55% female) were included (mean age in years (standard deviation): 61 (7.8)) at baseline (Figure S1). A total of 2,385 (incidence rate 4.0%) new PA-related cancer events occurred in the follow-up window (median follow up: (interquartile range (IQR)): 8.0 (7-9) years) and 1,552 participants died of other causes during follow-up. The included sample were predominantly of white ethnic backgrounds (96%). A large proportion had a college or university degree (43%), never smoked (56%) and consumed alcohol within guideline levels (57%) (Table 1).

**Table 1.** Included (n=59,218) UK Biobank participant characteristics at baseline accelerometry collection (June 2013-2015).

|  |  | Total Sample<br>n=59,218 |  |
| --- | --- | --- | --- |
| **Age (years)** | Mean, SD | 61.7 | 7.8 |
| **Sex (female)** | n, % | 32,452 | 55% |
| **Ethnicity (White)** | n, % | 56,988 | 96% |
| **Education** | n, % |  |  |
| College/University degree |  | 25,737 | 43% |
| A/AS-level |  | 7966 | 13% |
| NVQ/HND/HNC |  | 3235 | 6% |
| CSE |  | 2335 | 4% |
| O-levels |  | 12,116 | 21% |
| Other |  | 7829 | 13% |
| **Smoking history** | n, % |  |  |
| Current |  | 3925 | 7% |
| Never |  | 34,092 | 56% |
| Ex-smoker |  | 21,201 | 37% |
| **Medication Use** (Cholesterol lowering,<br>Blood pressure-lowering, Insulin) | n, % | 13,549 | 23% |
| **Alcohol** | n, % |  |  |
| Above guidelines |  | 22,409 | 38% |
| Ex-alcohol drinker |  | 1545 | 3% |
| Within guidelines |  | 33,647 | 57% |
| Never |  | 1617 | 3% |
| **Family History of Cancer** |  | 15,353 | 26% |
| **Diet Quality (Fruit/Vegetable**<br>**consumption) index** | Median (Q1,Q3) | 7 | (5-10) |
| **Previous History of CVD** |  | 5366 | 9% |
| **PA-related Cancer Incidence** |  | 2385 | 4% |
| **Follow up in years** | Median (Q1,Q3) | 8 | (7.4-8.5) |

Abbreviations: CVD: Cardiovascular disease; PA: Physical activity; NVQ: National Vocational Qualification; HND: Higher National Diploma; HNC: Higher National Certificate; CSE: Certificate of Secondary Education. Eligible sample included complete Physical activity measures, cancer outcomes and/or covariates (excluded sample presented in supplementary table S15).

### Participant movement compositions

The overall sample mean for composition 1 (posture) was 7h 39 min sleeping, 11h 13 min of ST, 4h 17 min standing, and 51 min moving (Figure S3). Participants who developed cancer spent 11.1% less time moving, 1.7% less time standing, 1.6% more ST compared with those without a cancer diagnosis, with no clear differences in sleep duration (Figure 1).

**Figure 1:**
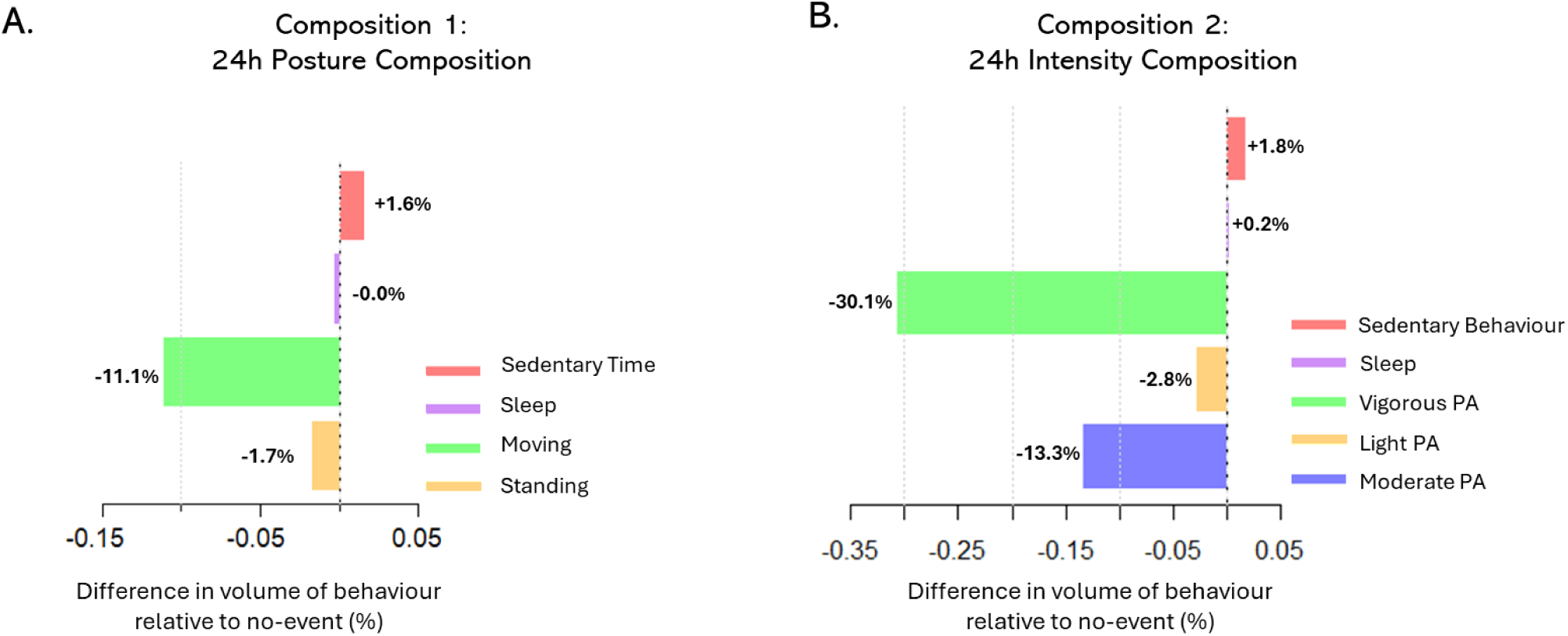
Proportional differences in movement (posture and intensity) and sleep compositions between participants with cancer vs no cancer. ***Legend:*** Proportional differences in the compositional average of movement behaviour, grouped by **A.** posture and **B.** intensity, of participants with incident PA-related cancer event in the follow-up window (n=2,385) relative to participants with no PA-related cancer event (n=55,281) over a median follow-up of 8.2 years.

The overall sample mean for composition 2 (intensity) was 7h 42m sleeping,10h 44m of SB, 5 hours (h) 3m of LPA, 28m of MPA, and 3m of VPA (Figure S2). Individuals with a cancer diagnosis spent 30.1%, 13.3% and 2.8% less time in VPA, MPA and LPA and 1.8% more time sedentary than those with no diagnosis. As above, there was no difference in sleep duration.

### Associations between Composition 1 (24-h posture) and cancer incidence

A greater proportion of time spent moving or standing, each relative to all other behaviours, was inversely associated with cancer incidence, while greater ST, relative to other behaviours was associated with greater cancer risk (Table S2). When examining all possible behavioural replacements, the greatest theoretical reduction in cancer risk from behavioural changes to the sample average for Composition 1 involved replacing sleep or ST with moving (Figure 2). For example, replacing 15 min of sleep or 15 min of ST with moving were each associated with HR of 0.98 (95% Confidence Interval: 0.97-0.99; Figure 2A-B, Tables S3-S5). However, replacing sleep or ST with standing required upwards of 30 min of replacement to observe equivalent risk reduction (30 min sleep replaced with 30 min standing, HR: 0.97, 95%CI: 0.96-0.99; 30 min ST replaced with 30 min Standing, HR: 0.98, 95%CI: 0.96-0.99; Figure 2C-D).

**Figure 2:**
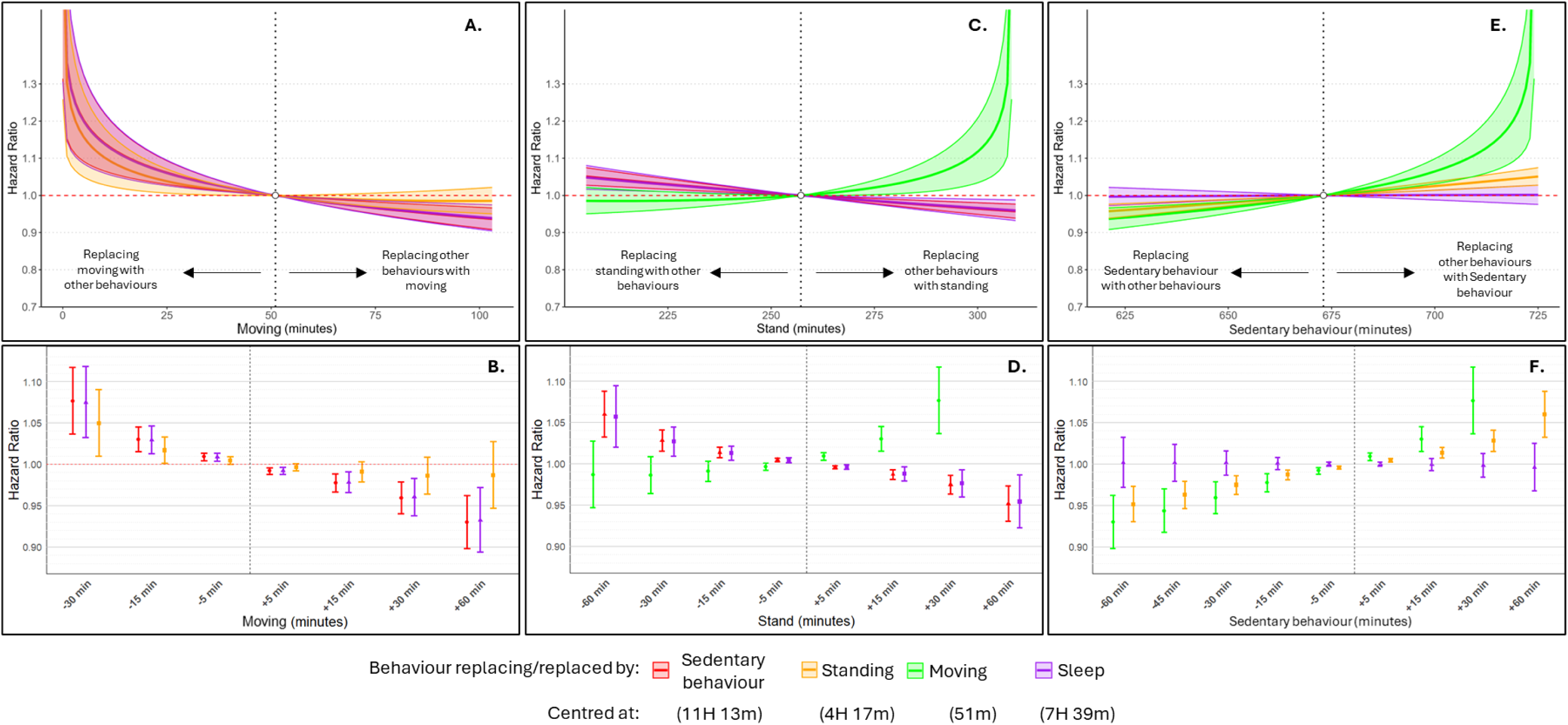
Difference in PA-related cancer hazard from theoretical reallocation of time between composition 1 (postures). ***Legend:*** Estimated difference in cancer hazard from theoretical reallocation of moving (**A,B**), standing (**C,D**) and sedentary behaviour (**E,F**) with/in place of each other postures and sleep, while holding each other behaviour constant around the sample average. Isotemporal substitution is performed on compositional Fine-Gray cox proportional hazards model, with 2,385 PA-related cancer events in n=59, 218; median 8.2y follow-up, and adjusted for age, sex, ethnicity, smoking status, alcohol use, diet quality, family history of cancer, history or presence of cardiovascular disease and regular medication use. Omitted bars are theoretical substitutions which exceed the range of the compositional data.

Conversely, compositions with less standing or moving and greater ST or sleep were associated with a greater cancer incidence (30 min moving replaced with 30 min ST, HR: 1.08, 95%CI: 1.04-1.12; 30 min moving replaced with 30 min sleep, HR: 1.07, 95%CI: 1.03-1.12; 30 min standing replaced with 30 min ST, HR: 1.03, 95%CI: 1.02-1.04; 30 min standing replaced with 30 min sleep, HR: 1.03, 95%CI: 1.01-1.04; Figure 2E-F).

### Associations between Composition 2 (24-h movement intensity) with cancer incidence

Greater time in VPA and LPA, each relative to all other behaviours, was inversely associated with cancer incidence (Table S6). Increasing daily VPA in place of other behaviours was associated with the greatest reduction in cancer risk (Figure 2, Tables S7,S8). For instance, 3 min additional VPA in place of any other intensity was associated with lower cancer risk (3 min sleep replaced with 3 min VPA, HR: 0.96, 95%CI: 0.93-0.99; 3 min SB replaced with 3 min VPA, HR: 0.96, 95%CI: 0.93-0.99; 3 min LPA replaced with 3 min VPA, HR: 0.96, 95%CI: 0.93-0.99; 3 min MPA replaced with 3 min VPA, HR: 0.96, 95%CI: 0.93-0.99). The only comparable risk reduction from changes to Composition 2 required increases in LPA in excess of 90 min, in place of sleep (HR: 0.92 (0.86-0.99)) or SB (HR: 0.92 (0.87-0.99)).

Similar to composition 1 (postures) reallocating time from VPA or LPA into SB or sleep was associated with increased cancer risk (Figure 3A-D). Results of all incremental time-reallocations are presented in Figures 1,2 & Tables S3-8.

**Figure 3:**
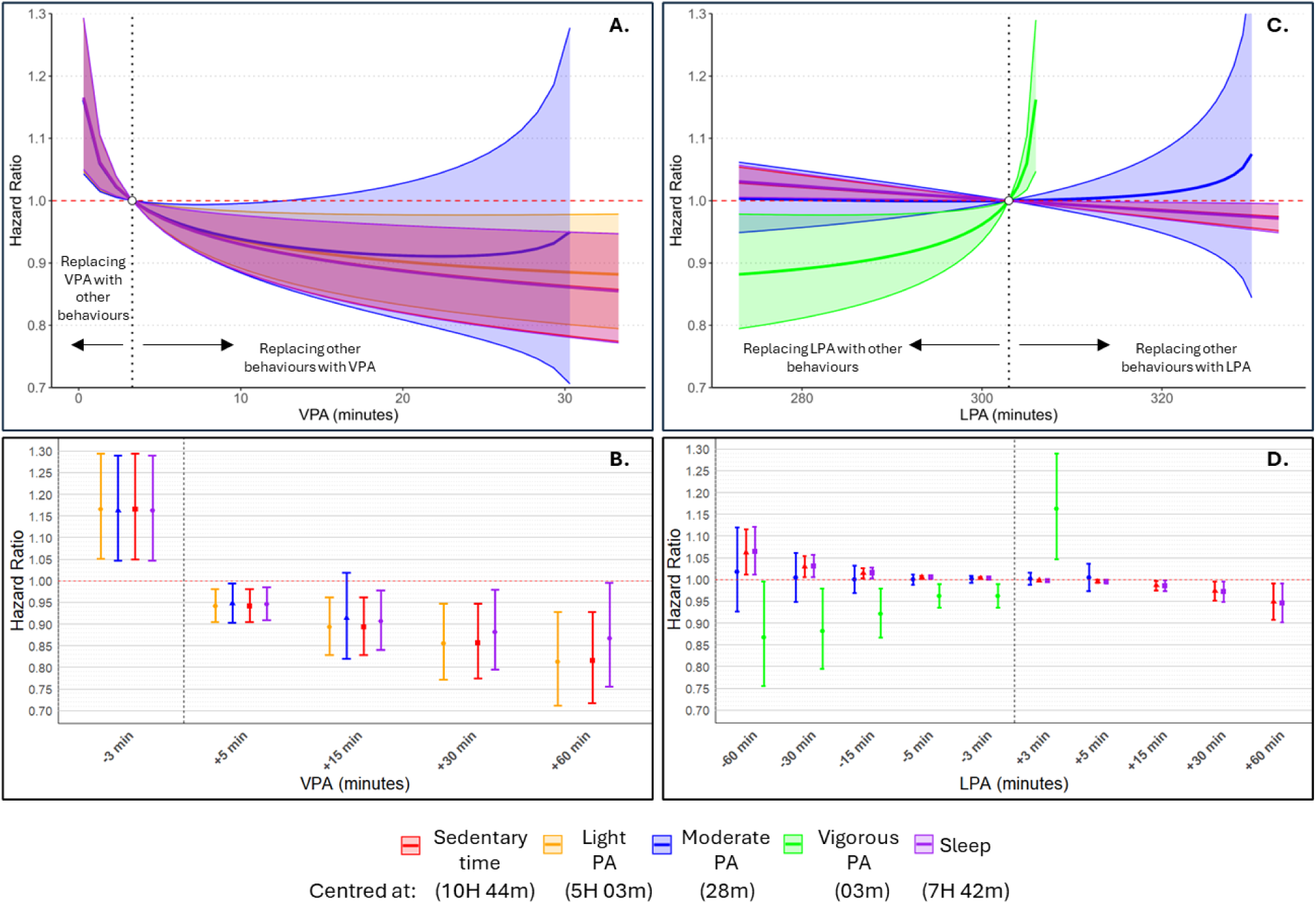
Difference in PA-related cancer hazard from theoretical reallocation of time between composition 2 (intensity). ***Legend:*** Estimated difference in PA-related cancer hazard from theoretical reallocation of vigorous physical activity (VPA) (**A,B**) and light physical activity (LPA) (**C,D**) into/in place of each other movement intensity and sleep, while holding each other behaviour constant around the sample average. Isotemporal substitution is performed on compositional Fine-Gray cox proportional hazards model, with 2,385 PA-related cancer events in n=59, 218; median 8.2y follow-up, and adjusted for age, sex, ethnicity, smoking status, family history of cancer, history or presence of cardiovascular disease, alcohol, diet quality, and regular medication use. Omitted bars are theoretical substitutions which exceed the range of the compositional data

Similar patterns were observed when re-fitted with MVPA. Greater time spent in MVPA, relative to other behaviours was inversely associated with cancer risk (Table S9). However, the odds of cancer incidence declined as the proportion of VPA within MVPA increased (Figure S3).

### Sensitivity analysis

E-values indicated that an unmeasured confounder must have an association of magnitude between 1.12-1.24, and 1.19-1.63 with both the exposure and outcome to nullify the weakest, and strongest reported associations respectively (Tables S10, S11). Removing events in the first (n=243 events) and second year (n=533 events) of follow-up did not alter the results (Tables S12-S15), however, associations between cancer risk and VPA were no longer significant after removing the first two years of events, despite no significant differences in the average compositions of these individuals from the original sample (Figures S4-6). Models involving MVPA proved robust to removal of the first two years of events (Table S15). Models involving all cancer sites showed only subtle deviation from the main analyses (Tables S16-17). The excluded sample were older, proportionally more female, and a greater proportion had a family history of cancer (Table S18).

## DISCUSSION

We adopted compositional data analysis methods to model the full spectrum of 24-h movement behaviours (categorised by posture and intensity) in relation to the incidence of PA-related cancer. When examining the posture composition, focused on the average participant’s day, more time (15 min) moving or more time standing—relative to all other postures—was associated with ∼1-7% lower cancer risk, while greater time spent sedentary was associated with higher cancer risk. When categorised by intensity, greater VPA and MVPA relative to other behaviours, followed by greater LPA relative to other behaviours, was associated with lower cancer risk, though there was less evidence of an association between MPA alone and cancer risk. As in the posture composition, greater time spent sedentary in place of VPA or LPA was associated with greater cancer risk in an exponential manner. Together, these findings extend our understanding of how risk of cancer may be associated with replacements of daily movement behaviour. They highlight how reallocation of sedentary time into more positive behaviours (e.g. standing, moving, and moving at a higher intensity) can decrease cancer risk, while the reverse (reallocation of moving or standing into SB) can increase risk of cancer. These findings may provide a broader range of options for recommendations for 24-h movement behaviour for mitigating cancer risk, enhancing applicability to the wider population.

Few large-scale studies have thusfar investigated the role of daily movement, including SB and sleep with risk of cancer^42, 43^. The extant literature has predominantly relied on self-reported PA^42^ and SB, or on SB proxies such as television-viewing^6, 7, 42^ and separately implicates leisure-time PA as protective, and SB as harmful^3, 42, 44^. One crucial device-based study extends this finding to daily behaviour, demonstrating the benefits of even short bursts of VPA throughout the 24-h day in non-exercisers for reducing cancer risk^36^, independently from other behaviours. Yet, only one device-based study has formally applied a 24-h movement framework in relation to cancer risk, revealing both greater MVPA and LPA in the 24-h day to be associated with reduced cancer risk^45^. Our study extends these findings, revealing nuances not previously reported, by examining both posture and intensity in a dual-compositional approach. For instance, we reveal how reallocating time to standing from sitting or sleep, may provide independent protective benefits, while VPA showed consistent inverse associations, even at short durations. Meanwhile, MPA alone showed no such association, unless combined with vigorous-intensity PA into MVPA. By incorporating both posture and intensity dimensions, and investigating these dimensions more deeply, this study provides the most detailed and nuanced exploration of 24-h movement behaviours and cancer risk to-date.

There are several hypothesized biologic mechanisms that may explain these findings t Some evidence exists that physical activity reduces chronic inflammation and insulin resistance while SB may increase these metabolic imbalances. ^46, 47^. Atypically, the less consistent finding for MPA alone may suggest that some combination with more intense movement is needed to trigger the physiological mechanisms that are critically related to cancer pathways^47 14^. LPA however, which is performed in larger volumes than MPA or VPA, may act through different mechanisms for cancer risk reduction, such as contributing more to overall daily movement and energy expenditure by minimising SB, which in all analyses appeared theoretically harmful. When grouped by posture, the potential mechanisms—in particular mechanisms linking standing with cancer—remain less clear^48^. We hypothesise that standing postures, as with LPA, may primarily act to break-up SB bouts, that increase cancer risk^6, 20^. The observed lack of associations of sleep with cancer align with the largest meta-analysis on this topic, which found limited evidence of an association with cancer for either long or short self-reported sleep duration^9^. Although beyond the scope of our study, it is possible that other dimensions of sleep (including quality measures and consistency) and the co-existence of sleep disorders (e.g. sleep apnoea), may still be mechanistically more important for shaping cancer risk, beyond duration alone^49^.

The above findings have substantial implications for public health policy. Where current movement guidelines focus exclusively on intensity and adopt a one-size-fits-all approach, we show how both changes in postural and intensity (e.g. increasing positive postures and more intensive movement), alongside reducing SB may act synergistically. Future research is needed to confirm if these findings could be the basis for behaviour change interventions for effective cancer prevention.

### Strengths and limitations

Strengths include the unique dual composition approach, and combination of contemporary approaches for the measurement, processing and modelling of 24-h movement data from accelerometers. Validated in free-living conditions, these procedures greatly enhanced the ability to capture dimensions of movement -posture and intensity– largely impossible using self-reported measures. Specifically, the use of compositional analysis permits the examination of all movement behaviours in tandem; a major limitation of prior studies that examine one or few behaviours, independently. Secondly, this study provides informative metrics on optimal dose to guide intervention efforts. Finally, procedures were used to account for multiple possible sources of bias, including reverse causation and quantification of residual confounding.

There were limitations including the small E-values leaving open the possibility for residual confounding nullifying findings smallest in magnitude. Possibility for misclassification of behaviours^16, 50^. The focus on the chosen cancer sites was based on self-reported measures of LTPA^3^, of which there is incomplete consensus^42^, and other cancer types may still show associations with other facets of daily movement, including sleep. Finally, the UK Biobank study has known healthy-volunteer bias, reducing its generalisability to the wider UK population^51^. However, previous empirical evidence suggests that the poor representativeness of the UK Biobank did not materially affect the associations of PA with cancer outcomes^52^.

## CONCLUSIONS

Greater time spent either standing or moving in place of SB and sleep was inversely associated with cancer risk. Alternatively, when defined by intensity, VPA or MVPA, in place of any other behaviours, or LPA in place of sleep or SB were associated with lower cancer risk. These findings provide a range of possible alternative options for drawing recommendations for movement behaviours for cancer risk reduction, which may be accessible to a larger proportion of and the population to aid cancer prevention.

## Abbreviations

PA: Physical activity
VPA: Vigorous physical activity
LPA: Light physical activity
MPA: Moderate physical activity
SB: Sedentary behaviour
ST: Sedentary time
h: Hour
min: Minute
WHO: World Health Organization
CVD: cardiovascular disease

## DECLARATIONS

### Data availability

The data that support the findings of this study are available from the UK Biobank but restrictions apply to the availability of these data, which were used under license for the current study, and so are not publicly available.

### Author Contributions

Mitchell, Biswas and Ahmadi had access to the data in the study and conducted the data analysis.

Concept and design: Mitchell, Stamatakis, Ahmadi, Koemel, Hamer.

Acquisition, analysis, or interpretation of data: Mitchell, Biswas, Ahmadi, Blodgett.

Drafting of the manuscript: Mitchell.

Critical revision of the manuscript for important intellectual content: All authors.

Statistical analysis: Mitchell, Biswas, Koemel, Ahmadi

Obtained funding: Stamatakis, Hamer.

Administrative, technical, or material support: Stamatakis, Ahmadi.

Supervision: Stamatakis, Hamer.

## Supplementary materials

Supplementary material_V9.1.docx

