## Supplementary material_V9.1 for "Device-measured movement behaviours and cancer incidence in a population sample of UK adults: Dual 24-hour analyses of postural and intensity compositions"

### Supplementary Text S1. Classification of intensity and posture from processing algorithms.

#### Sleep classifier

Sleep duration was assessed with an algorithm, using the absence of change in device tilt angle, developed and validated in 3,752 British participants with sleep diary data and 28 patients with polysomnography data. The sleep period time window derived from the algorithm was 11m and 3m longer compared with a sleep diary in men and women, respectively. The mean c-statistic to detect the sleep period time window compared to polysomnography was 0.86 and 0.83 in clinic-based and healthy sleepers, respectively[1].

#### Physical activity intensity classifier

Physical activity intensity was classified with a previously adopted two-level Random Forest algorithm[2, 3]. The first level classifies activity type, in contiguous 10s bins, and the second level further sorts the activities into intensity bands. This two-level classifier minimises misclassification between the lower intensity bands as activities are first classified by type, clearly delineating Moving activities, standing utilitarian movements (semi-stationary) and sedentary behaviour (SB) before non-sedentary activities are further classified into intensity bands based on degree of acceleration against gravity. Moving activities are then grouped by their acceleration, as light PA (LPA; <100mg), moderate PA (MPA; ≥100mg) and vigorous (≥400mg). The intensity composition therefore encompassed VPA, MPA, LPA, sedentary time (ST) and sleep.

#### Posture classifier

Stationary behaviour and physical activity were classified using a previously validated Random Forest (RF) activity classifier. The initial probabilities of activity classification from the RF classifier were temporally smoothed using a Hidden Markov Model (HMM) to generate the final sequence of activity predictions.

Physical activity was categorized in 60-second intervals into five classes: Sitting, Standing Stationary, Standing Moving, Walking/Running, and Cycling and has been shown to have a high degree of classification accuracy[6, 374, 5]. Sleep and non-wear time were distinguished from active periods based on changes in tilt angle and acceleration standard deviation. Accelerometer data were calibrated and orientation-corrected according to established procedures (model specificity and accuracy metrics have been previously outlined by Ahmadi et al., 2024).

*Matthew N Ahmadi, Pieter Coenen, Leon Straker, Emmanuel Stamatakis, Device-measured stationary behaviour and cardiovascular and orthostatic circulatory disease incidence, International Journal of Epidemiology, Volume 53, Issue 6, December 2024, dyae136,* [*https://doi.org/10.1093/ije/dyae136*](https://doi.org/10.1093/ije/dyae136)

### Supplementary Text S2. Isometric log-ratio *'Pivot'* approach to Compositional data analysis.

The pivot approach[6, 7] to compositional data analysis is established for enabling the modelling of highly collinear movement behaviours in tandem. This involves applying the Isometric log-ratio transformation to compositional data of *n* components, resulting in *n-1* coordinates which represent all daily movement behaviour in a series of ratios, and which can all be fitted within standard statistical models. This circumvents issues of multicollinearity[6].

The first ILR-coordinate within each model represents the association of one movement behaviour, relative to all others (i.e. VPA relative to MPA, LPA, SB and sleep) with the outcome, and is the only interpreted and reported value in each model. The first ILR coefficeint represents the expected difference in the outcome when the ILR-coordinate is increased by one unit. The order of the ratios is then re-pivoted, and the model re-fitted, to investigate the association of a different component, relative to all others, with the outcome.

Figure S1. Sample derivation flow chart

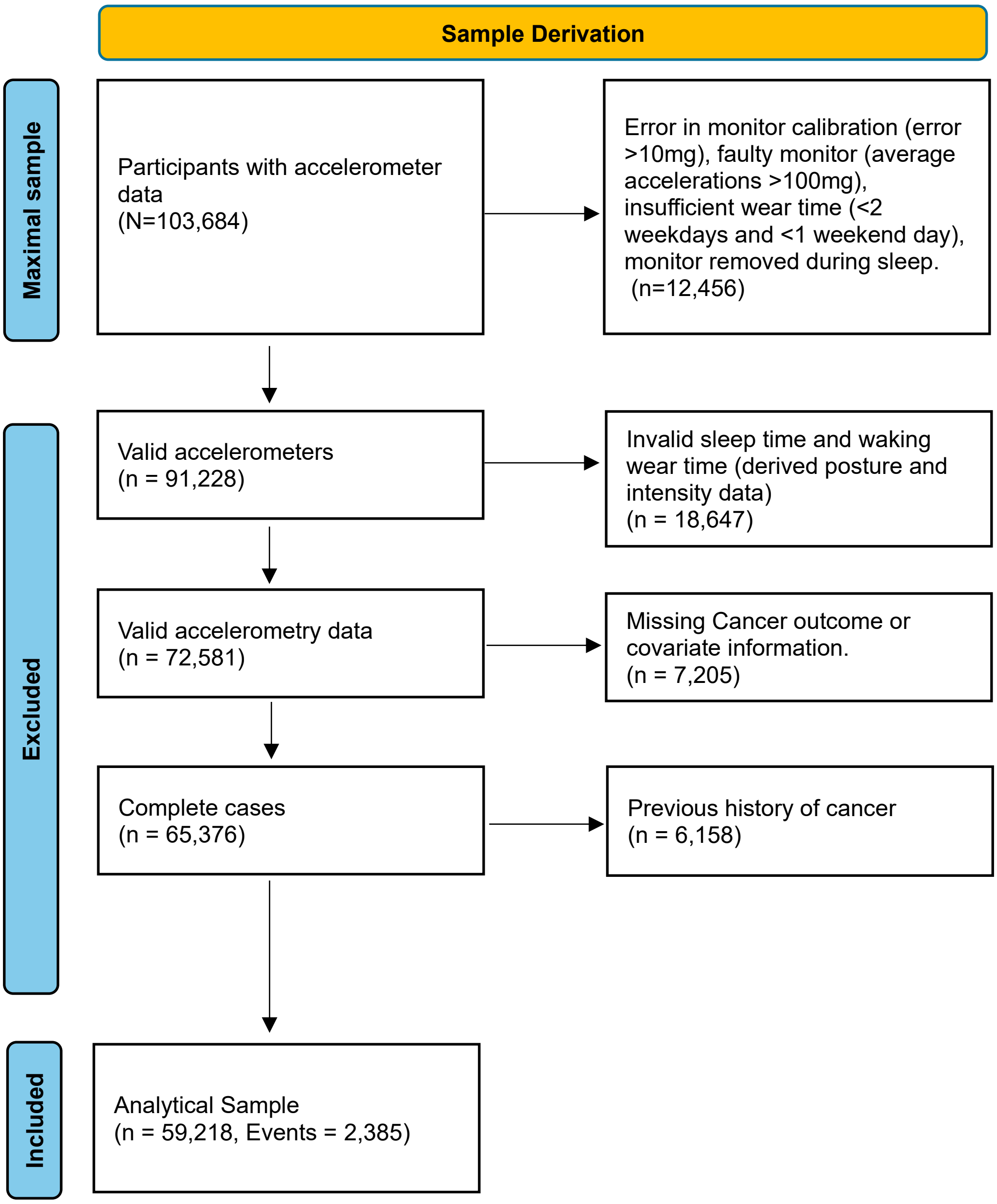

Figure S2. Raw sample means of (left: composition 1 (Posture); right: composition 2 (Intensity)

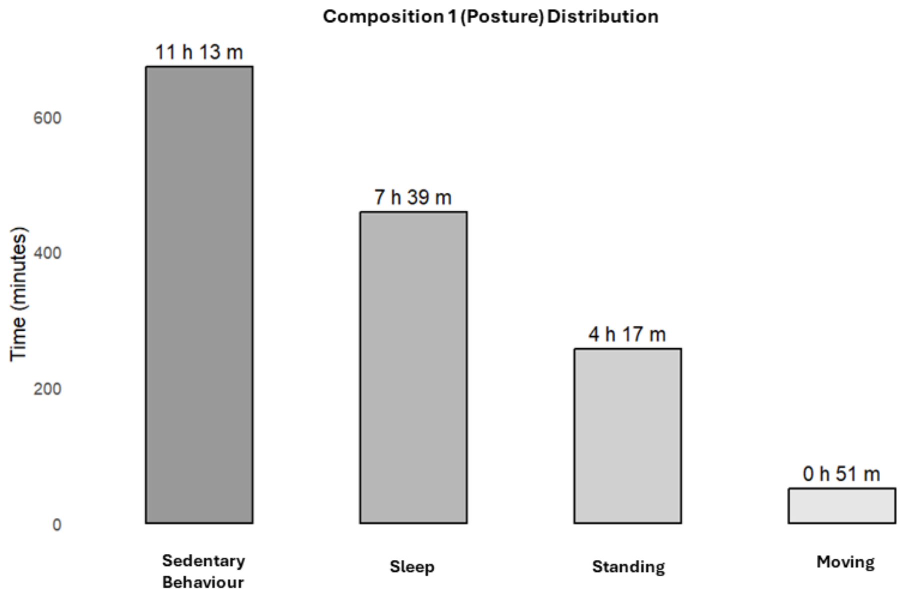


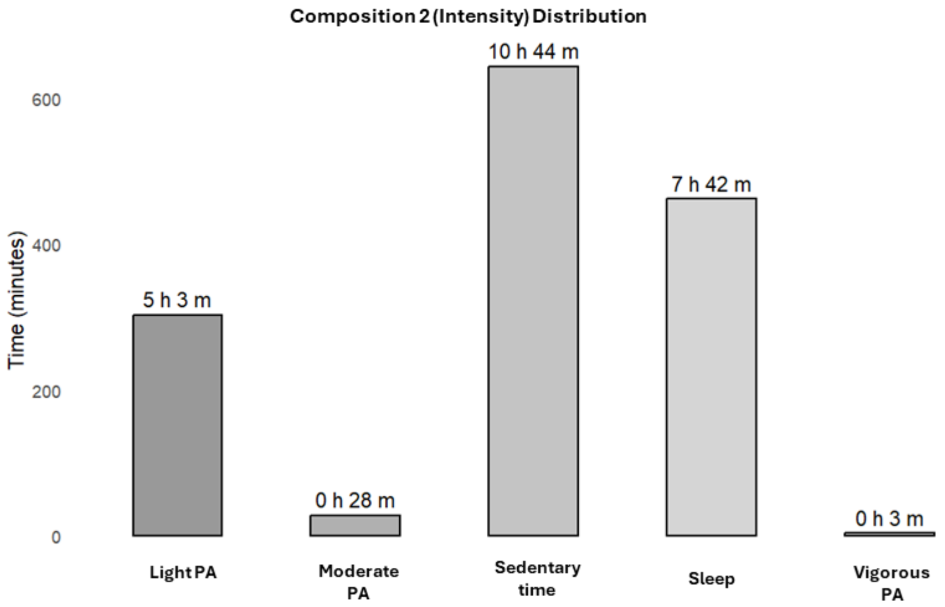

| **Table S1. Placeholder Table ICD Codes / Biobank Var numbers** | | |
|---|---|---|
| Variable | Coding | UKB field |
| Age | Continuous (years) | 34, 52, accelerometer date-timestamp |
| Sex | Female/Male | 31 |
| Smoker status | Never, past, current smoker. | 20116 |
| Alcohol Use | Never drinker, Ex-drinker Within guidelines and 4 units/wk, Above guidelines and | 20117, 1558 |
| Diet | Fruits and vegetables servings/day | 1309, 1319, 1289, 1299 |
| Age at first PA-related cancer event | Physical activity-related cancer at 13 sites (esophageal adenocarcinoma, liver, lung, kidney, gastric cardia, endometrial, myeloid leukemia, myeloma, colon, head and neck, rectal, bladder, and breast) using ICD-10 classifications: C0-C3, C5-C9, C40-C42, C45-C49. | 100092 |
| Prevalent cancer | | 20001, 100092 |
| Prevalent CVD | Yes/No Identified by self-report and hospitalisation (ICD-10: I0, I11, I13, I20-I51, I60-I69). | 20002, 41259 |

| | | |
|---|---|---|
| Education | College/University; A/AS level; O levels; CSE; NVQ/HND/HNC; other. A/AS = Advanced Placement; O level = High school certificate; CSE = Certificate of secondary education; NVQ/HND/HNC = Vocational qualification / Associates degree. A level, typically at age 18 years; O level, typically at age 16 years; CSE, typically at age 16 years; | 6138 |
| cholesterol medication | Yes/No | 6177, 6153 |
| blood pressure medication | Yes/No | 6177, 6153 |
| Use of diabetes medication | Yes/No | 6177, 6153 |
| Parental history of cancer | Yes/No Self-reported mother or father diagnosed with cancer | 20107, 20110 |

**Table S2. Association of Coordinates of composition 1 (posture) and sleep with risk of PA-related cancers (Fine-Gray Cox Proportional Hazards Model)**

| | **Age and Sex-adjusted** | | | | **Fully adjusted** | | | |
|---|---|---|---|---|---|---|---|---|
| | **HR** | **95% CI** | | **p-value** | **HR** | **95% CI** | | **p-value** |
| **Moving** (vs all other postures) | **0.957** | **0.937** | **0.978** | **<0.001** | **0.962** | **0.942** | **0.983** | **<0.001** |
| **Standing** (vs all other postures) | **0.911** | **0.872** | **0.953** | **<0.001** | **0.917** | **0.877** | **0.959** | **<0.001** |
| **Sedentary Behaviour** (vs all other postures) | **1.095** | **1.026** | **1.169** | **0.006** | **1.084** | **1.016** | **1.158** | **0.015** |
| Sleep (vs all other postures) | 1.047 | 0.969 | 1.131 | 0.241 | 1.045 | 0.968 | 1.129 | 0.259 |

**Magnitude of Coefficients are ILR transformed units and not directly interpretable.**

**Table S3 Predicted HR from Isotemporal substitution around sample average of Moving whilst holding other components constant at sample average.**

| | MOVING→SEDENTARY BEHAVIOUR | | | MOVING→STANDING | | | MOVING→SLEEP | | |
|---|---|---|---|---|---|---|---|---|---|
| | HR | 95% CI | | HR | 95% CI | | HR | 95% CI | |
| **-90 min** | **-** | - | - | - | - | - | **-** | - | - |
| **-60 min** | **-** | - | - | - | - | - | **-** | - | - |
| **-45 min** | **1.19** | 1.08 | 1.30 | **1.14** | 1.04 | 1.25 | **1.18** | 1.08 | 1.30 |
| **-30 min** | **1.08** | 1.04 | 1.12 | **1.05** | 1.01 | 1.09 | **1.07** | 1.03 | 1.12 |
| **-15 min** | **1.03** | 1.01 | 1.05 | **1.02** | 1.00 | 1.03 | **1.03** | 1.01 | 1.05 |
| **-10 min** | **1.02** | 1.01 | 1.03 | **1.01** | 1.00 | 1.02 | **1.02** | 1.01 | 1.03 |
| **-5 min** | **1.01** | 1.00 | 1.01 | 1.00 | 1.00 | 1.01 | **1.01** | 1.00 | 1.01 |
| **-3 min** | **1.01** | 1.00 | 1.01 | 1.00 | 1.00 | 1.01 | **1.01** | 1.00 | 1.01 |
| | **SEDENTARY BEHAVIOUR→MOVING** | | | **STANDING→MOVING** | | | **SLEEP→MOVING** | | |
| **+3 min** | **1.00** | 0.99 | 1.00 | 1.00 | 1.00 | 1.00 | **1.00** | 0.99 | 1.00 |
| **+5 min** | **0.99** | 0.99 | 1.00 | 1.00 | 0.99 | 1.00 | **0.99** | 0.99 | 1.00 |
| **+10 min** | **0.98** | 0.98 | 0.99 | 0.99 | 0.98 | 1.00 | **0.98** | 0.98 | 0.99 |
| **+15 min** | **0.98** | 0.97 | 0.99 | 0.99 | 0.98 | 1.00 | **0.98** | 0.97 | 0.99 |
| **+30 min** | **0.96** | 0.94 | 0.98 | 0.99 | 0.96 | 1.01 | **0.96** | 0.94 | 0.98 |
| **+45 min** | **0.94** | 0.92 | 0.97 | 0.98 | 0.95 | 1.02 | **0.95** | 0.91 | 0.98 |
| **+60 min** | **0.93** | 0.90 | 0.96 | 0.99 | 0.95 | 1.03 | **0.93** | 0.89 | 0.97 |
| **+90 min** | **0.91** | 0.87 | 0.95 | 1.00 | 0.94 | 1.06 | **0.91** | 0.86 | 0.96 |

**Table S4 Predicted HR from Isotemporal substitution around sample average of standing whilst holding other components constant at sample average.**

| | MOVING→STANDING | | | STANDING→SEDENTARY BEHAVIOUR | | | STANDING→SLEEP | | |
|---|---|---|---|---|---|---|---|---|---|
| | HR | 95% CI | | HR | 95% CI | | HR | 95% CI | |
| **-90 min** | **-** | - | - | 1.10 | 1.05 | 1.14 | **1.09** | 1.03 | 1.15 |
| **-60 min** | **-** | - | - | 1.06 | 1.03 | 1.09 | **1.06** | 1.02 | 1.09 |
| **-45 min** | **1.14** | 1.04 | 1.25 | **1.04** | 1.02 | 1.06 | **1.04** | 1.01 | 1.07 |
| **-30 min** | **1.05** | 1.01 | 1.09 | **1.03** | 1.02 | 1.04 | **1.03** | 1.01 | 1.04 |
| **-15 min** | **1.02** | 1.00 | 1.03 | **1.01** | 1.01 | 1.02 | **1.01** | 1.00 | 1.02 |
| **-10 min** | **1.01** | 1.00 | 1.02 | **1.01** | 1.00 | 1.01 | **1.01** | 1.00 | 1.01 |
| **-5 min** | 1.00 | 1.00 | 1.01 | 1.00 | 1.00 | 1.01 | **1.00** | 1.00 | 1.01 |
| **-3 min** | 1.00 | 1.00 | 1.01 | 1.00 | 1.00 | 1.00 | **1.00** | 1.00 | 1.00 |
| | **STANDING→MOVING** | | | **SEDENTARY BEHAVIOUR→STANDING** | | | **SLEEP→STANDING** | | |
| **+3 min** | 1.00 | 1.00 | 1.00 | **1.00** | 1.00 | 1.00 | **1.00** | 1.00 | 1.00 |
| **+5 min** | 1.00 | 0.99 | 1.00 | **1.00** | 0.99 | 1.00 | **1.00** | 0.99 | 1.00 |
| **+10 min** | 0.99 | 0.98 | 1.00 | **0.99** | 0.99 | 1.00 | **0.99** | 0.99 | 1.00 |
| **+15 min** | 0.99 | 0.98 | 1.00 | **0.99** | 0.98 | 0.99 | **0.99** | 0.98 | 1.00 |
| **+30 min** | 0.99 | 0.96 | 1.01 | **0.97** | 0.96 | 0.99 | **0.98** | 0.96 | 0.99 |
| **+45 min** | 0.98 | 0.95 | 1.02 | **0.96** | 0.95 | 0.98 | **0.96** | 0.94 | 0.99 |
| **+60 min** | 0.99 | 0.95 | 1.03 | **0.95** | 0.93 | 0.97 | **0.95** | 0.92 | 0.99 |
| **+90 min** | 1.00 | 0.94 | 1.06 | **0.93** | 0.90 | 0.96 | **0.93** | 0.89 | 0.98 |

**Table S5 Predicted HR from Isotemporal substitution around sample average of Sedentary Behaviour postures whilst holding other components constant at sample average.**

| | MOVING→SEDENTARY BEHAVIOUR | | | STANDING→SEDENTARY BEHAVIOUR | | | SLEEP→SEDENTARY BEHAVIOUR | | |
|---|---|---|---|---|---|---|---|---|---|
| | HR | 95% CI | | HR | 95% CI | | HR | 95% CI | |
| **-90 min** | **-** | - | - | 1.1 | 1.05 | 1.14 | 1 | 0.96 | 1.05 |
| **-60 min** | **-** | - | - | 1.06 | 1.03 | 1.09 | 1 | 0.97 | 1.03 |
| **-45 min** | **1.19** | 1.08 | 1.3 | **1.04** | 1.02 | 1.06 | 1 | 0.98 | 1.02 |
| **-30 min** | **1.08** | 1.04 | 1.12 | **1.03** | 1.02 | 1.04 | 1 | 0.99 | 1.02 |
| **-15 min** | **1.03** | 1.01 | 1.05 | **1.01** | 1.01 | 1.02 | 1 | 0.99 | 1.01 |
| **-10 min** | **1.02** | 1.01 | 1.03 | **1.01** | 1 | 1.01 | 1 | 1 | 1.01 |
| **-5 min** | **1.01** | 1 | 1.01 | **1** | 1 | 1.01 | 1 | 1 | 1 |
| **-3 min** | **1.01** | 1 | 1.01 | **1** | 1 | 1 | 1 | 1 | 1 |
| | **SEDENTARY BEHAVIOUR→MOVING** | | | **SEDENTARY BEHAVIOUR→STANDING** | | | **SEDENTARY BEHAVIOUR→SLEEP** | | |
| **+3 min** | **1** | 0.99 | 1 | 1 | 1 | 1 | 1 | 1 | 1 |
| **+5 min** | **0.99** | 0.99 | 1 | 1 | 0.99 | 1 | 1 | 1 | 1 |
| **+10 min** | **0.98** | 0.98 | 0.99 | **0.99** | 0.99 | 1 | 1 | 0.99 | 1 |
| **+15 min** | **0.98** | 0.97 | 0.99 | **0.99** | 0.98 | 0.99 | 1 | 0.99 | 1.01 |
| **+30 min** | **0.96** | 0.94 | 0.98 | **0.97** | 0.96 | 0.99 | 1 | 0.98 | 1.01 |
| **+45 min** | **0.94** | 0.92 | 0.97 | **0.96** | 0.95 | 0.98 | 1 | 0.98 | 1.02 |
| **+60 min** | **0.93** | 0.9 | 0.96 | **0.95** | 0.93 | 0.97 | 1 | 0.97 | 1.02 |
| **+90 min** | **0.91** | 0.87 | 0.95 | **0.93** | 0.9 | 0.96 | 0.99 | 0.95 | 1.04 |

| **Table S6. Association of Coordinates of composition 2 (intensity) and sleep with risk of PA-related cancers (Fine-Gray Cox Proportional Hazards Model)** | | | | | | | | |
|---|---|---|---|---|---|---|---|---|
| | **Age and Sex-adjusted** | | | | **Fully adjusted** | | | |
| | **HR** | **95% CI** | | **p-value** | **HR** | **95% CI** | | **p-value** |
| **Vigorous PA** (vs all other behaviours) | **0.922** | **0.878** | **0.968** | **0.001** | **0.932** | **0.887** | **0.978** | **0.004** |
| Moderate PA (vs all other behaviours) | 0.971 | 0.903 | 1.043 | 0.415 | 0.973 | 0.905 | 1.046 | 0.454 |
| **Light PA** (vs all other behaviours) | **0.781** | **0.642** | **0.950** | **0.013** | **0.792** | **0.651** | **0.965** | **0.020** |
| Sleep (vs all other behaviours) | 1.169 | 0.948 | 1.442 | 0.145 | 1.165 | 0.945 | 1.437 | 0.152 |
| Sedentary time (vs all other behaviours) | 1.225 | 0.993 | 1.510 | 0.058 | 1.195 | 0.969 | 1.475 | 0.096 |
| **Magnitude of Coefficients are ILR transformed units and not directly interpretable.** | | | | | | | | |

**Table S7. Predicted HR from Isotemporal substitution around sample average of VPA whilst holding other components constant at sample average.**

| **Exchanging VPA→** | **→ST** | | | **→MPA** | | | **→SLEEP** | | | **→LPA** | | |
|---|---|---|---|---|---|---|---|---|---|---|---|---|
| | HR | 95% CI | | HR | 95% CI | | HR | 95% CI | | HR | 95% CI | |
| **-3 min** | **1.17** | 1.05 | 1.29 | **1.16** | 1.05 | 1.29 | **1.17** | 1.05 | 1.29 | **1.16** | 1.05 | 1.29 |
| **+3 min** | **0.96** | 0.93 | 0.99 | **0.96** | 0.93 | 0.99 | **0.96** | 0.93 | 0.99 | **0.96** | 0.93 | 0.99 |
| **+5 min** | **0.94** | 0.90 | 0.98 | **0.95** | 0.90 | 0.99 | **0.94** | 0.90 | 0.98 | **0.95** | 0.91 | 0.99 |
| **+10 min** | **0.91** | 0.86 | 0.97 | 0.92 | 0.85 | 1.00 | **0.91** | 0.86 | 0.97 | **0.92** | 0.87 | 0.98 |
| **+15 min** | **0.89** | 0.83 | 0.96 | 0.91 | 0.82 | 1.02 | **0.89** | 0.83 | 0.96 | **0.91** | 0.84 | 0.98 |
| **+30 min** | **0.86** | 0.77 | 0.95 | | | | **0.85** | 0.77 | 0.95 | **0.88** | 0.79 | 0.98 |
| **+45 min** | **0.83** | 0.74 | 0.94 | | | | **0.83** | 0.74 | 0.94 | **0.87** | 0.77 | 0.99 |
| **+60 min** | **0.82** | 0.72 | 0.93 | | | | **0.81** | 0.71 | 0.93 | **0.87** | 0.76 | 1.00 |
| **+90 min** | **0.79** | 0.68 | 0.91 | | | | **0.78** | 0.67 | 0.91 | 0.87 | 0.74 | 1.02 |

**Table S8. Predicted HR from Isotemporal substitution around sample average of LPA whilst holding other components constant at sample average.**

| **Exchange LPA→** | **→ST** | | | **→MPA** | | | **→SLEEP** | | | **→VPA** | | |
|---|---|---|---|---|---|---|---|---|---|---|---|---|
| | HR | 95% CI | | HR | 95% CI | | HR | 95% CI | | HR | 95% CI | |
| **-90 min** | **1.10** | 1.02 | 1.19 | 1.04 | 0.91 | 1.18 | **1.10** | 1.02 | 1.19 | **0.87** | 0.74 | 1.02 |
| **-60 min** | **1.06** | 1.01 | 1.12 | 1.02 | 0.93 | 1.12 | **1.06** | 1.01 | 1.12 | **0.87** | 0.76 | 1.00 |
| **-45 min** | **1.05** | 1.01 | 1.08 | 1.01 | 0.94 | 1.09 | **1.05** | 1.01 | 1.09 | **0.87** | 0.77 | 0.99 |
| **-30 min** | **1.03** | 1.01 | 1.05 | 1.00 | 0.95 | 1.06 | **1.03** | 1.01 | 1.06 | **0.88** | 0.79 | 0.98 |
| **-15 min** | **1.01** | 1.00 | 1.03 | 1.00 | 0.97 | 1.03 | **1.02** | 1.00 | 1.03 | **0.92** | 0.87 | 0.98 |
| **-10 min** | **1.01** | 1.00 | 1.02 | 1.00 | 0.98 | 1.02 | **1.01** | 1.00 | 1.02 | **0.95** | 0.91 | 0.99 |
| **-5 min** | **1.00** | 1.00 | 1.01 | 1.00 | 0.99 | 1.01 | **1.00** | 1.00 | 1.01 | **0.96** | 0.93 | 0.99 |
| **-3 min** | **1.00** | 1.00 | 1.01 | 1.00 | 0.99 | 1.01 | **1.00** | 1.00 | 1.01 | **0.96** | 0.93 | 0.99 |
| **+3 min** | **1.00** | 0.99 | 1.00 | 1.00 | 0.99 | 1.02 | **1.00** | 0.99 | 1.00 | **1.16** | 1.05 | 1.29 |
| **+5 min** | **1.00** | 0.99 | 1.00 | 1.00 | 0.97 | 1.04 | **1.00** | 0.99 | 1.00 | | | |
| **+10 min** | **0.99** | 0.98 | 1.00 | 1.01 | 0.96 | 1.06 | **0.99** | 0.98 | 1.00 | | | |
| **+15 min** | **0.99** | 0.98 | 1.00 | | | | **0.99** | 0.97 | 1.00 | | | |
| **+30 min** | **0.97** | 0.95 | 0.99 | | | | **0.97** | 0.95 | 1.00 | | | |
| **+45 min** | **0.96** | 0.93 | 0.99 | | | | **0.96** | 0.92 | 0.99 | | | |
| **+60 min** | **0.95** | 0.91 | 0.99 | | | | **0.94** | 0.90 | 0.99 | | | |
| **+90 min** | **0.92** | 0.87 | 0.99 | | | | **0.92** | 0.86 | 0.99 | | | |

| Table S9 Association of Coordinates of daily-movement behaviours (intensities, including MVPA) and sleep with risk of PA-related cancers (Fine-Gray Cox Proportional Hazards Model) (including all events). | | | | | | | | |
|---|---|---|---|---|---|---|---|---|
| | **Age and Sex-adjusted** | | | | **Fully adjusted** | | | |
| | **HR** | **95% CI** | | **p-value** | **HR** | **95% CI** | | **p-value** |
| **Moderate-to-Vigorous PA** (vs all other behaviours) | **0.950** | **0.923** | **0.979** | **0.001** | **0.956** | **0.928** | **0.985** | **0.003** |
| **Light PA** (vs all other behaviours) | **0.891** | **0.813** | **0.975** | **0.012** | **0.897** | **0.819** | **0.983** | **0.019** |
| Sleep (vs all other behaviours) | 1.073 | 0.974 | 1.182 | 0.155 | 1.072 | 0.973 | 1.181 | 0.162 |
| Sedentary time (vs all other behaviours) | 1.101 | 0.999 | 1.213 | 0.052 | 1.088 | 0.987 | 1.199 | 0.090 |
| **Coefficients are ILR transformed units and not directly interpretable.** | | | | | | | | |

| **Table S10. E-values of posture model** | | |
|---|---|---|
| Exposure | E-value | (Lower-limit) |
| Moving (vs other postures) | 1.242 | (1.151) |
| Standing (vs other postures) | 1.318 | (1.204) |
| Sedentary Behaviour (vs other postures) | 1.305 | (1.116) |
| Sleep (vs other postures) | Null | - |

Point estimate (and upper limit of the confidence interval in brackets) that an unmeasured confounder must have with both the exposure and outcome (conditional on the measured covariates) to render the exposure-outcome association null.

| **Table S11. E-values of Intensity model** | | |
|---|---|---|
| Exposure | E-value | (Lower-limit) |
| Vigorous PA (vs other movement behaviours) | 1.282 | (1.143) |
| Moderate PA (vs other movement behaviours) | Null | - |
| Light PA (vs other movement behaviours) | 1.628 | (1.187) |
| Sedentary time (vs other movement behaviours) | Null | - |
| Sleep (vs other movement behaviours) | Null | - |

Point estimate (and upper limit of the confidence interval in brackets) that an unmeasured confounder must have with both the exposure and outcome (conditional on the measured covariates) to render the exposure-outcome association null.

**Table S12. Association of Coordinates of daily-movement behaviours (postures) and sleep with risk of PA-related cancers (Fine-Gray Cox Proportional Hazards Model) with removal of first 1y follow-up.**

| | **Age and Sex-adjusted** | | | | **Fully adjusted** | | | |
|---|---|---|---|---|---|---|---|---|
| | **HR** | **95% CI** | | **p-value** | **HR** | **95% CI** | | **p-value** |
| **Moving** (vs all other postures) | **0.960** | **0.939** | **0.981** | **<0.001** | **1.034** | **0.941** | **0.987** | **0.002** |
| **Standing** (vs all other postures) | **0.917** | **0.875** | **0.961** | **<0.001** | **0.923** | **0.881** | **0.968** | **<0.001** |
| **Sedentary Behaviour** (vs all other postures) | 1.088 | 1.015 | 1.164 | 0.017 | 1.075 | 1.003 | 1.151 | 0.040 |
| Sleep (vs all other postures) | 1.045 | 0.961 | 1.134 | 0.285 | 1.044 | 0.963 | 1.132 | 0.300 |

**Magnitude of Coefficients are ILR transformed units and not directly interpretable.**

**Table S13. Association of Coordinates of daily-movement behaviours (intensities) and sleep with risk of PA-related cancers (Fine-Gray Cox Proportional Hazards Model) with removal of first 1y follow-up.**

| | Age and Sex-adjusted | | | | Fully adjusted | | | |
|---|---|---|---|---|---|---|---|---|
| | **HR** | **95% CI** | | **p-value** | **HR** | **95% CI** | | **p-value** |
| **Vigorous PA** (vs all other behaviours) | **0.936** | **0.890** | **0.985** | **0.018** | **0.946** | **0.899** | **0.997** | **0.037** |
| Moderate PA (vs all other behaviours) | 0.959 | 0.889 | 1.035 | 0.277 | 0.962 | 0.891 | 1.038 | 0.319 |
| **Light PA** (vs all other behaviours) | 1.156 | 0.927 | 1.443 | 0.199 | 1.125 | 0.901 | 1.405 | 0.299 |
| Sleep (vs all other behaviours) | **0.795** | **0.795** | **0.647** | **0.030** | **0.807** | **0.807** | **0.656** | **0.042** |
| Sedentary time (vs all other behaviours) | 1.212 | 0.969 | 1.513 | 0.091 | 1.210 | 0.969 | 1.511 | 0.093 |

**Magnitude of Coefficients are ILR transformed units and not directly interpretable.**

**Table S14. Association of Coordinates of daily-movement behaviours (postures) and sleep with risk of PA-related cancers (Fine-Gray Cox Proportional Hazards Model) with removal of first 2y follow-up.**

| | **Age and Sex-adjusted** | | | | **Fully adjusted** | | | |
|---|---|---|---|---|---|---|---|---|
| | **HR** | **95% CI** | | **p-value** | **HR** | **95% CI** | | **p-value** |
| **Moving** (vs all other postures) | **0.954** | **0.932** | **0.978** | **0.000** | **0.960** | **0.937** | **0.984** | **0.001** |
| **Standing** (vs all other postures) | **0.920** | **0.875** | **0.967** | **0.001** | **0.926** | **0.880** | **0.974** | **0.003** |
| **Sedentary Behaviour** (vs all other postures) | 1.088 | 1.010 | 1.172 | 0.026 | 1.084 | 1.016 | 1.158 | 0.015 |
| Sleep (vs all other postures) | 1.047 | 0.969 | 1.042 | 0.134 | 1.044 | 0.957 | 1.140 | 0.331 |

**Magnitude of Coefficients are ILR transformed units and not directly interpretable.**

**Table S15. Association of Coordinates of daily-movement behaviours (intensities) and sleep with risk of PA-related cancers (Fine-Gray Cox Proportional Hazards Model) with removal of first 2y follow-up.**

| | Age and Sex-adjusted | | | | Fully adjusted | | | |
|---|---|---|---|---|---|---|---|---|
| | HR | 95% CI | | p-value | HR | 95% CI | | p-value |
| Vigorous PA (vs all other behaviours) | 0.948 | 0.897 | 1.001 | 0.056 | 0.958 | 0.906 | 1.012 | 0.126 |
| Moderate PA (vs all other behaviours) | 0.937 | 0.863 | 1.016 | 0.116 | 0.940 | 0.866 | 1.020 | 0.135 |
| **Light PA** (vs all other behaviours) | 0.759 | 0.607 | 0.949 | 0.015 | 0.771 | 0.616 | 0.963 | 0.022 |
| Sleep (vs all other behaviours) | 1.226 | 0.965 | 1.557 | 0.096 | 1.220 | 0.961 | 1.550 | 0.102 |
| Sedentary time (vs all other behaviours) | 1.211 | 0.954 | 1.537 | 0.116 | 1.182 | 0.930 | 1.501 | 0.172 |

**Magnitude of Coefficients are ILR transformed units and not directly interpretable.**

**Figure S6** Association of ratio variables (proportion of VPA and MPA within MVPA) with risk of PA-related cancers (Age, sex, MVPA and SB-adjusted Fine-Gray Cox Proportional Hazards Model, with knots placed at the 10th, 50th and, 90th percentile).

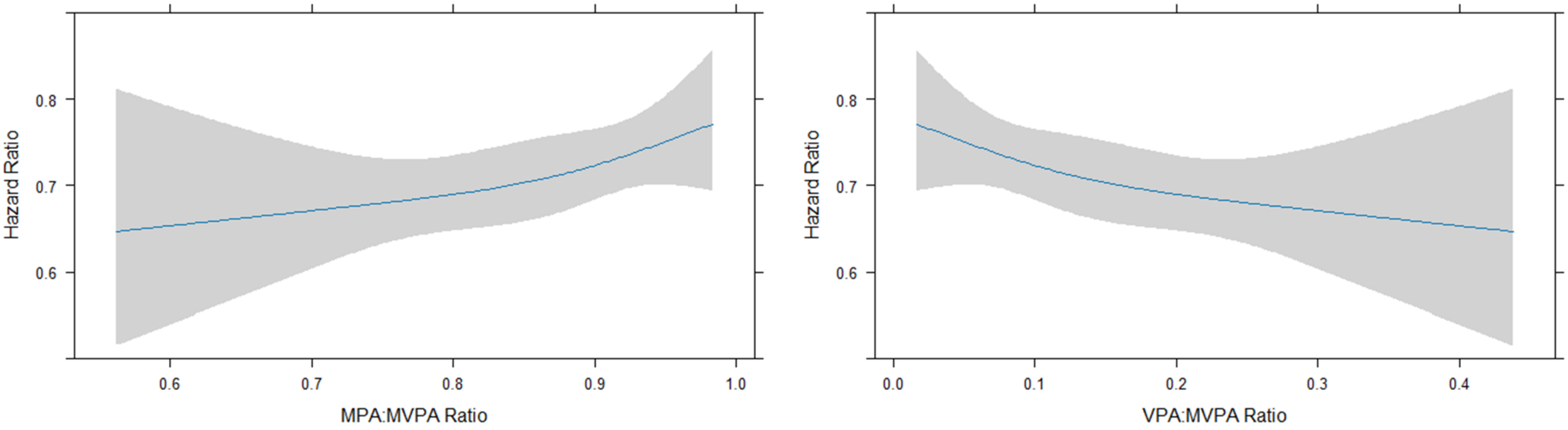


**Adjusted to:** MVPA= 34m; SB=10h 36m; Sex=Male; Age=62y

**Adjusted to:** MVPA= 34m; SB=10h 36m; Sex=Male; Age=62y

**Figure S3**. Raw sample means of composition 2 (intensity) (PA-related Cancer incidence within first 2y follow-up)

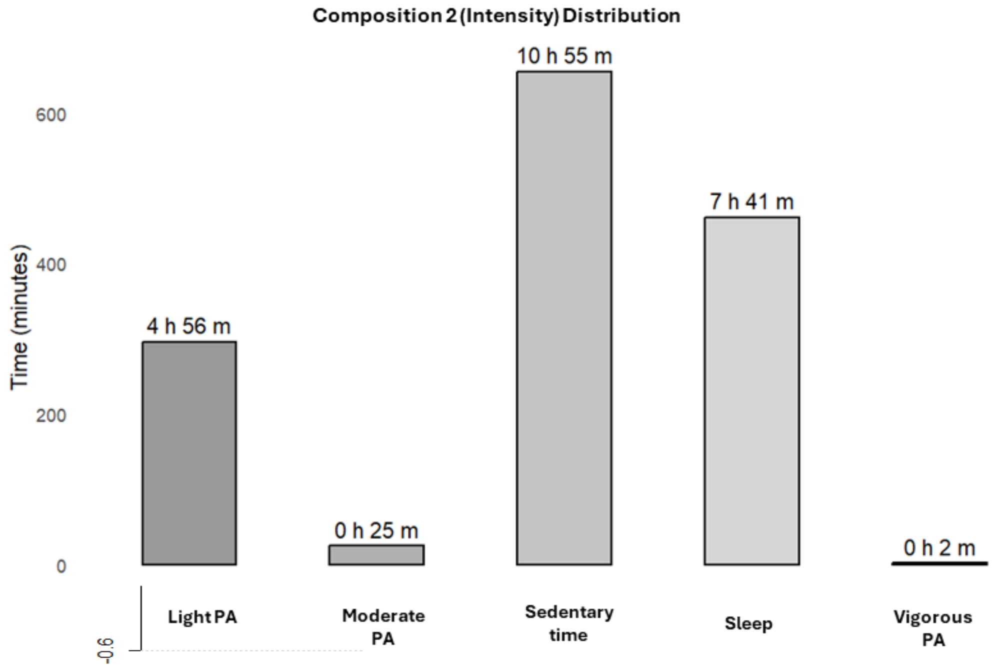


**Figure S4**. Difference in composition 2 (intensity) of individuals with PA-related Cancer incidence within first 1y follow-up, relative to sample average.

| Vigorous PA | Sleep | Moderate PA | Light PA | Sedentary time |
|---|---|---|---|---|
| 2.12 | 460.18 | 23.2 | 294.41 | 660.1 |

**Figure S5**. Difference in composition 2 (intensity) of individuals with PA-related Cancer incidence excluding first 2y follow-up, relative to sample average.

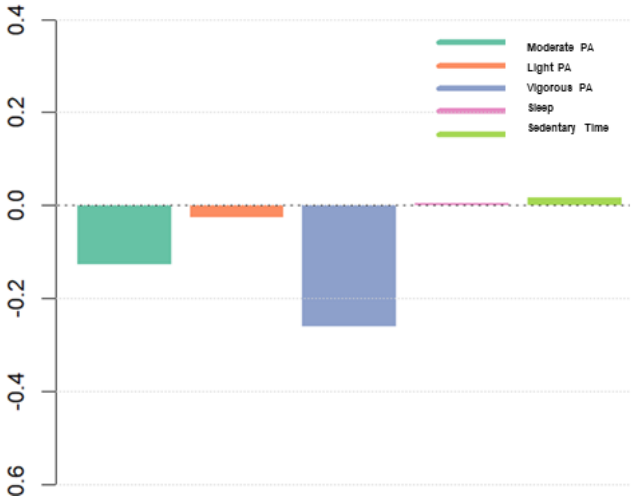


| Vigorous PA* | Sleep | Moderate PA | Light PA | Sedentary time |
|---|---|---|---|---|
| 2.56 | 463.82 | 24.5 | 295.1 | 654.06 |

MANOVA Pillai's trace test of multivariate differences in compositional centre between PA-related cancer in first 2y follow-up compared to outside of 2y follow-up (p=0.2405)

*Significant t-test on difference in compositional average VPA between PA-related cancer in first 2y follow-up compared to outside of 2y follow-up (p<0.0177).

| Table S15. Association of Coordinates of daily-movement behaviours (intensities, including MVPA) and sleep with risk of PA-related cancers (Fine-Gray Cox Proportional Hazards Model) (after removal of first 2y of events). | | | | | | | | |
|---|---|---|---|---|---|---|---|---|
| | **Age and Sex-adjusted** | | | | **Fully adjusted** | | | |
| | **HR** | **95% CI** | | **p-value** | **HR** | **95% CI** | | **p-value** |
| **Moderate-to-Vigorous PA** (vs all other behaviours) | **0.946** | **0.914** | **0.977** | **0.001** | **0.952** | **0.92** | **0.985** | **0.0038** |
| **Light PA** (vs all other behaviours) | **0.881** | **0.772** | **1.006** | **0.0159** | **0.888** | **0.805** | **0.979** | **0.0232** |
| Sleep (vs all other behaviours) | 1.09 | 0.98 | 1.22 | 0.1139 | 1.08 | 0.97 | 1.21 | 0.1719 |
| Sedentary time (vs all other behaviours) | 1.098 | 0.973 | 1.241 | 0.0967 | 1.096 | 0.983 | 1.211 | 0.1028 |
| **Coefficients are ILR transformed units and not directly interpretable.** | | | | | | | | |

| **Table S16. Association of Coordinates of daily-movement behaviours (postures) and sleep with risk of All cancer sites (Fine-Gray Cox Proportional Hazards Model) with removal of first 2y follow-up.** | | | | | | | | |
|---|---|---|---|---|---|---|---|---|
| | **Age and Sex-adjusted** | | | | **Fully adjusted** | | | |
| | **HR** | **95% CI** | | **p-value** | **HR** | **95% CI** | | **p-value** |
| **Moving** (vs all other postures) | 0.97 | 0.96 | 0.98 | <0.001 | 0.97 | 0.96 | 0.99 | <0.001 |
| **Standing** (vs all other postures) | 0.96 | 0.94 | 0.99 | 0.004 | 0.96 | 0.94 | 0.99 | 0.009 |
| Sedentary Behaviour (vs all other postures) | 1.00 | 0.96 | 1.04 | 0.924 | 1.00 | 0.96 | 1.04 | 0.951 |
| **Sleep** (vs all other postures) | 1.07 | 1.02 | 1.12 | 0.004 | 1.07 | 1.02 | 1.12 | 0.007 |
| **Magnitude of Coefficients are ILR transformed units and not directly interpretable.** | | | | | | | | |

| **Table S17. Association of Coordinates of daily-movement behaviours (intensities) and sleep with risk of All cancer sites (Fine-Gray Cox Proportional Hazards Model) with removal of first 2y follow-up.** | | | | | | | | |
|---|---|---|---|---|---|---|---|---|
| | **Age and Sex-adjusted** | | | | **Fully adjusted** | | | |
| | **HR** | **95% CI** | | **p-value** | **HR** | **95% CI** | | **p-value** |
| Vigorous PA (vs all other behaviours) | 0.98 | 0.95 | 1.01 | 0.170 | 0.98 | 0.95 | 1.01 | 0.265 |
| Moderate PA (vs all other behaviours) | 0.98 | 0.94 | 1.02 | 0.386 | 0.98 | 0.94 | 1.02 | 0.401 |
| **Light PA** (vs all other behaviours) | 0.85 | 0.76 | 0.96 | 0.008 | 0.86 | 0.76 | 0.96 | 0.009 |
| **Sleep** (vs all other behaviours) | 1.22 | 1.07 | 1.38 | 0.002 | 1.20 | 1.06 | 1.36 | 0.005 |
| Sedentary time (vs all other behaviours) | 1.00 | 0.89 | 1.14 | 0.945 | 1.01 | 0.89 | 1.15 | 0.861 |
| **Magnitude of Coefficients are ILR transformed units and not directly interpretable.** | | | | | | | | |

**Table S18. Included and excluded participant characteristics at baseline.**

| | | Included Sample n=59,218 | | Excluded Sample n=22,666 | | p-value |
|---|---|---|---|---|---|---|
| **Age (years)** | Mean (SD) | 61.7 | (7.8) | 62.9 | (7.8) | **<0.001** |
| **Sex (female)** | n, % | 32452 | 55% | 13549 | 60% | **<0.001** |
| **Ethnicity (White)** | n, % | 56988 | 96% | 21632 | 95% | **<0.001** |
| **Education** | n, % | | | | | **0.007** |
| College/University degree | | 25737 | 43% | 9997 | 44% | |
| A/AS-level | | 7966 | 13% | 2943 | 13% | |
| NVQ/HND/HNC | | 3235 | 6% | 1151 | 5% | |
| CSE | | 2335 | 4% | 832 | 4% | |
| O-levels | | 12116 | 21% | 4622 | 20% | |
| Other | | 7829 | 13% | 3121 | 14% | |
| **Smoking history** | n, % | | | | | **<0.001** |
| Current | | 3925 | 7% | 1633 | 7% | |
| Never | | 34092 | 56% | 12621 | 56% | |
| Ex | | 21201 | 37% | 8412 | 37% | |
| **Medication Use** (Cholesterol lowering, Blood pressure-lowering, Insulin) | n, % | 13549 | 23% | 5687 | 25% | **<0.001** |
| **Alcohol** | n, % | | | | | **<0.001** |
| Above guidelines | | 22409 | 38% | 8423 | 37% | |
| Ex | | 1545 | 3% | 627 | 3% | |
| Within guidelines | | 33647 | 57% | 12944 | 57% | |
| Never | | 1617 | 3% | 672 | 3% | |
| **Family history of Cancer** | | 15353 | 26% | 6424 | 29% | **<0.001** |
| **Diet Quality (Fruit/Vegetable consumption) index** | Median (Q1,Q3) | 7 | (5-10) | 7 | (5-10) | **0.1953** |
| **Previous history of CVD** | | 5366 | 9% | 2372 | 10% | **<0.001** |
| **PA-related Cancer Incidence** | | 2385 | 4% | - | - | - |
| **Follow up in years** | Median (Q1,Q3) | 8.0 | (7.4-8.5) | - | - | - |
| **Competing risk** | Median (Q1,Q3) | 1552 | 3% | - | - | - |
| **Previous history of Cancer** | | - | - | 7633 | 34% | - |

Comparisons of categorical variables with chi-squared tests and continuous variables with Wilcoxon signed-rank tests. Totals in excluded sample may vary due to missingness. (CVD: Cardiovascular disease; PA: Physical activity). Excluded sample were missing Physical activity measures, cancer outcomes and/or covariates.
